# Azithromycin Treatment Response as a Probe to Attribute Bacterial Aetiologies of Diarrhoea using Molecular Diagnostics: A Reanalysis of the AntiBiotics for Children with severe Diarrhoea (ABCD) Trial

**DOI:** 10.1101/2024.09.03.24312730

**Authors:** Jennifer Cornick, Sarah Elwood, James Platts-Mills, Patricia Pavlinac, Karim Manji, Chris Sudfeld, Christopher P. Duggan, Queen Dube, Naor Bar-Zeev, Karen Kotloff, Samba O Sow, Sunil Sazawal, Benson O Singa, Judd L Walson, Farah Qamar, Tahmeed Ahmed, Ayesha De Costa, Elizabeth T Rogawski McQuade

## Abstract

**Background:** Multi-pathogen molecular diagnostics enable assignment of diarrhoea aetiology, but defining thresholds of pathogen quantity to accurately attribute aetiology is challenging in high-burden settings where coinfections are common. The Antibiotics for Children with severe Diarrhoea (ABCD) trial provides an opportunity to leverage the azithromycin treatment response to inform which diarrhoea episodes are bacterial.

**Methods:** We analysed data from ABCD, which randomized children with watery diarrhoea to azithromycin or placebo. We quantified heterogeneity in the azithromycin treatment response by the quantity of enteric pathogens detected by qPCR as a tool for understanding aetiology.

**Results:** The heterogeneity in azithromycin treatment response was most prominent for *Shigella*. The risk ratio for diarrhoea on day 3 post enrolment for azithromycin compared to placebo was 13% (95% CI:3, 23) lower per log10 increase in *Shigella* quantity. The protective effect of azithromycin on diarrhoea at day 3 also became stronger as pathogen quantities increased for *Vibrio cholerae*, ST-ETEC, and tEPEC. No association between pathogen quantity and azithromycin response was observed for *Campylobacter*, LT-ETEC or EAEC. The associations were consistent for the outcome of 90-day hospitalisation or death.

**Conclusions:** The relationships between response to azithromycin treatment and bacterial pathogen quantities observed for *Shigella*, *Vibrio cholerae*, ST-ETEC and tEPEC confirm prior evidence that these pathogens are the likely cause of diarrhoea when detected at high quantities. The lack of a similar response pattern for *Campylobacter*, LT-ETEC or EAEC is consistent with the limited association between pathogen quantity and diarrhoea symptoms previously observed in large studies of diarrhoea aetiology.

****Key message** (3 succinct bullet points, each a single sentence):**

1. We investigated whether heterogeneity in treatment response observed in the ABCD trial, where children with diarrhoea were randomised to receive azithromycin or placebo, could be used to inform aetiological attribution of diarrhoea to bacterial enteric pathogens.
2. The protective effect of azithromycin on diarrhoea at day 3 and hospitalisation or death at day 90 became stronger as pathogen quantities increased for *Shigella, Vibrio cholerae, ST-ETEC and tEPEC* but not for *Campylobacter*, LT-ETEC or EAEC.
3. The relationships between *Shigella, Vibrio cholerae*, ST-ETEC and tEPEC quantity and response to antibiotic treatment confirm prior evidence that these pathogens are the likely cause of diarrhoea when detected at high quantities and could be used to inform which diarrhoea cases should be treated with antibiotics.

## Introduction

Diarrhoeal diseases are the second leading cause of death in children under five, with the highest burden of disease experienced in low- and middle- income countries (LMICs) (1). Accurate estimates of the burden of specific aetiologies of diarrhoea are necessary for global resource allocation and policy making and may be useful to inform appropriate treatment measures beyond the WHO IMCI recommended management (2). Quantitative PCR (qPCR) diagnostics have been used to attribute likely diarrhoea aetiologies in multiple large global studies, including the Global Enteric Multicentre Study (GEMS) (3), the Malnutrition and the Consequences for Child Health and Development (MAL-ED) study (4), and the Global Paediatric Diarrhoea Surveillance network (5).

The Antibiotics for Children with severe Diarrhoea (ABCD) study, a 7-country, randomized, double-blinded, placebo-controlled trial assessed if a 3-day course of azithromycin reduced mortality or improved linear growth among children with acute watery diarrhoea accompanied by dehydration or undernutrition (6). Rotavirus (21.1%) was the leading cause of diarrhoea, while 28.3% of diarrhoeal cases had a likely bacterial aetiology, most commonly *Shigella,* ST-ETEC, and typical EPEC (7). While the impact of azithromycin among all enrolled children was minimal, molecular diagnostics revealed that azithromycin was effective at reducing risk of day 3 diarrhoea and day 90 hospitalization or death among children with likely bacterial diarrhoea (7). Due to the absence of diarrhoea-free controls in ABCD, pathogen-specific quantity cut-offs were applied to assign etiology, based on the strong associations of those quantities with diarrhoea previously observed in MAL-ED and GEMS (3,4). However, there was a residual benefit of azithromycin among episodes with bacteria detected at a lower quantity. For example, children with a likely bacterial aetiology had a 3.1% absolute reduction in risk of 90-day hospitalization or death when treated with azithromycin compared to a 2.3% reduction among children with bacteria detected but not attributed (7). These findings suggest the previously used cut-offs likely missed true bacterial episodes that also responded to treatment.

The ABCD study provides a unique opportunity to further explore diarrhoeal aetiology based on azithromycin treatment response. Azithromycin is a broad-spectrum antibiotic with known efficacy against Gram-negative pathogens, including ETEC (8) *Campylobacter* spp (9) and *Shigella* spp (10,11). We therefore expect an azithromycin treatment response in many cases of diarrhoea in which bacteria are the true aetiological agent. We investigated if heterogeneity in the treatment response by pathogen quantity can further refine aetiological quantity cut-offs for enteric bacteria leading to more precise aetiologic assignment of diarrhoeal episodes.

## Methods

The ABCD study design (12), primary analysis results (6), and post-hoc analyses incorporating qPCR enteric pathogen testing (13) have been previously described. Briefly, children 2-23 months of age were enrolled between June 2017 and July 2019 from seven sites in Bangladesh, India, Kenya, Malawi, Mali, Pakistan, and Tanzania if they presented to study hospitals with acute watery diarrhoea. To be included, children had to have some or severe dehydration and/or moderate wasting or severe stunting, as previously defined (12) and could not have received antibiotics in the 2 weeks prior to presentation. Children with clear indications for antibiotic treatment (i.e., dysentery, severe acute malnutrition, signs of other infections requiring antibiotics) were excluded.

Enrolled children were randomized to receive either three days of azithromycin (10 mg/kg/day) or placebo. Before randomisation, study staff collected whole stool or a flocked rectal swab which was tested for enteric pathogens by qPCR for the first approximately 1,000 children enrolled at each site, using the TaqMan array card platform as described previously (12). To account for differences in sample type, cycle thresholds (Ct’s) derived from rectal swabs were adjusted by the pathogen-specific mean cycle threshold difference between paired rectal swabs and whole stools. Follow-up visits were conducted to ascertain presence of diarrhoea on day 3 after enrolment and rehospitalisation and vital status 90 days after enrolment.

Pathogens were analysed using the continuous Ct as a marker of relative pathogen quantity. To define co-aetiologies (i.e., coinfections at a quantity associated with diarrhoea), we used the pathogen-specific qPCR Ct cut-offs derived for ABCD (7) from two large multisite studies of diarrhoea that included non-diarrhoeal controls, GEMS (3) and MAL-ED (4). A co-aetiology was defined as the detection of any other pathogen at a quantity above (or equally, a qPCR Ct below) the aetiological cut-off. To maximize power to detect treatment effect heterogeneity and to be consistent with prior analyses (7), the primary outcome assessed in this analysis was presence of diarrhoea 3 days after enrolment. As a secondary outcome, we considered a combined outcome of rehospitalisation or death by 90 days after enrolment.

For each pathogen previously associated with diarrhoea (3) or with at least 5% prevalence, we modelled the heterogeneity in the azithromycin treatment effect by pathogen quantity detected using log-binomial regression adjusting for the day of diarrhoea on which the sample was collected and co-detection of other pathogens. Co-detection of other pathogens was specified with two variables representing the sum of the episode-specific attributable fractions for all bacteria and the sum of the episode-specific attributable fractions for all viruses and protozoa. Episode-specific attributable fractions (AFes) were derived using attribution models from GEMS and MAL-ED by applying pathogen quantities from ABCD diarrhoea episodes and taking the median of 1000 estimates drawn equally from each of the site-specific models (5). We first included an interaction term between treatment arm assignment and the quantity of pathogen detected when specified with a linear and quadratic term. We conducted a linear trend test for each interaction using the Wald-based p-value for the quadratic interaction term with alpha = 0.05. Because the test was not statistically significant for 13 of 15 pathogens evaluated (Table S1), we proceeded with modelling the interactions with only linear terms for pathogen quantity across all pathogens for interpretability and comparability. For the diarrhoea on day 3 outcome, we further stratified the models by presence of a bacterial co-aetiology.

We report the model-predicted pathogen quantity and treatment arm-specific risk of each outcome conditional on sample collection at day 0 and no co-detections using ggpredict from the ggeffects package v. 1.2.1 in R v. 4.2.1. We report the risk ratio for azithromycin compared to placebo for each outcome by pathogen quantity using the interplot 0.2.3 package, both overall and stratified by presence of a bacterial co- aetiology for the diarrhoea on day 3 outcome. These data were summarized by reporting the change in the effect of azithromycin per each log10 increase in pathogen quantity based on the interaction term between pathogen quantity and azithromycin. To estimate the proportion of episodes that could be attributed to each bacterial pathogen based on the azithromycin treatment response, we calculated population attributable fractions from these episode-specific risk ratios for day 3 diarrhea. Specifically, to remove azithromycin effects unrelated to the pathogen detected, we divided each episode-specific risk ratio by the largest risk ratio when pathogen quantity was zero (RR_adj_). The population attributable fraction was then calculated as 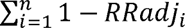 (since azithromycin is protective) where *i* included all episodes in which the pathogen was detected at any quantity. Adjusted risk ratios were constrained to be less than or equal to 1 such that the episode attributable fraction could not be negative.

Finally, across the range of cycle threshold values, we calculated the proportion of diarrhoea episodes that would be treated with antibiotics if a given pathogen-specific cycle threshold were chosen as the cut-off to define aetiology and assign treatment. This proportion was calculated among diarrhoea episodes overall to quantify the proportion of all diarrhoea episodes that would be treated using a given cut-off and among diarrhoea episodes with the pathogen detected at any quantity to quantify the proportion of all potentially pathogen-attributable episodes that would be treated.

## Results

We included 6692 children who were among the first 1000 enrolled at each of the seven ABCD trial sites and had their samples tested by qPCR with valid a result. This represents 80.9% of all children enrolled in the ABCD trial (n=8268). The density of quantities detected for each pathogen was similar between azithromycin and placebo groups (Figure S1).

The overall risk of diarrhoea on day 3 was 10.2% and was higher in the placebo (12.0%) compared to azithromycin (8.4%) group. For a subset of the enteric bacteria, specifically *Shigella,* ST-ETEC, tEPEC, and *Vibrio cholerae*, azithromycin was more strongly protective for diarrhoea on day 3 as pathogen quantity detected increased. For these pathogens, the risk of diarrhoea on day 3 in the placebo group increased with increasing pathogen quantity, while the risk in the azithromycin group was less dependent on pathogen quantity (Figure 1A). Correspondingly, the effect of azithromycin became stronger (i.e., risk ratios further from the null) as pathogen quantities increased (Figure 1B). The heterogeneity in azithromycin effect by pathogen quantity was most striking and statistically significant only for *Shigella.* The risk ratio for diarrhoea on day 3 for azithromycin compared to placebo was 13% (95% CI: 3, 23) lower (i.e., further from the null) per log10 increase in *Shigella* quantity detected (Table 1). This translated to detection at a high quantity (Ct = 25) associated with a 50% reduction (95% CI: 30, 64) in diarrhoea on day 3 compared to a 40% reduction (95% CI: 33, 75) associated with detection at Ct = 30. A similar pattern was observed for *V. cholerae,* ST-ETEC, and tEPEC, though the effect heterogeneity was not statistically significant for these pathogens. When stratifying by whether there was a bacterial co- aetiology, the association between pathogen quantity and azithromycin treatment response was slightly stronger for episodes in which there was no co-aetiology (Figure 1C).

**Figure 1.**
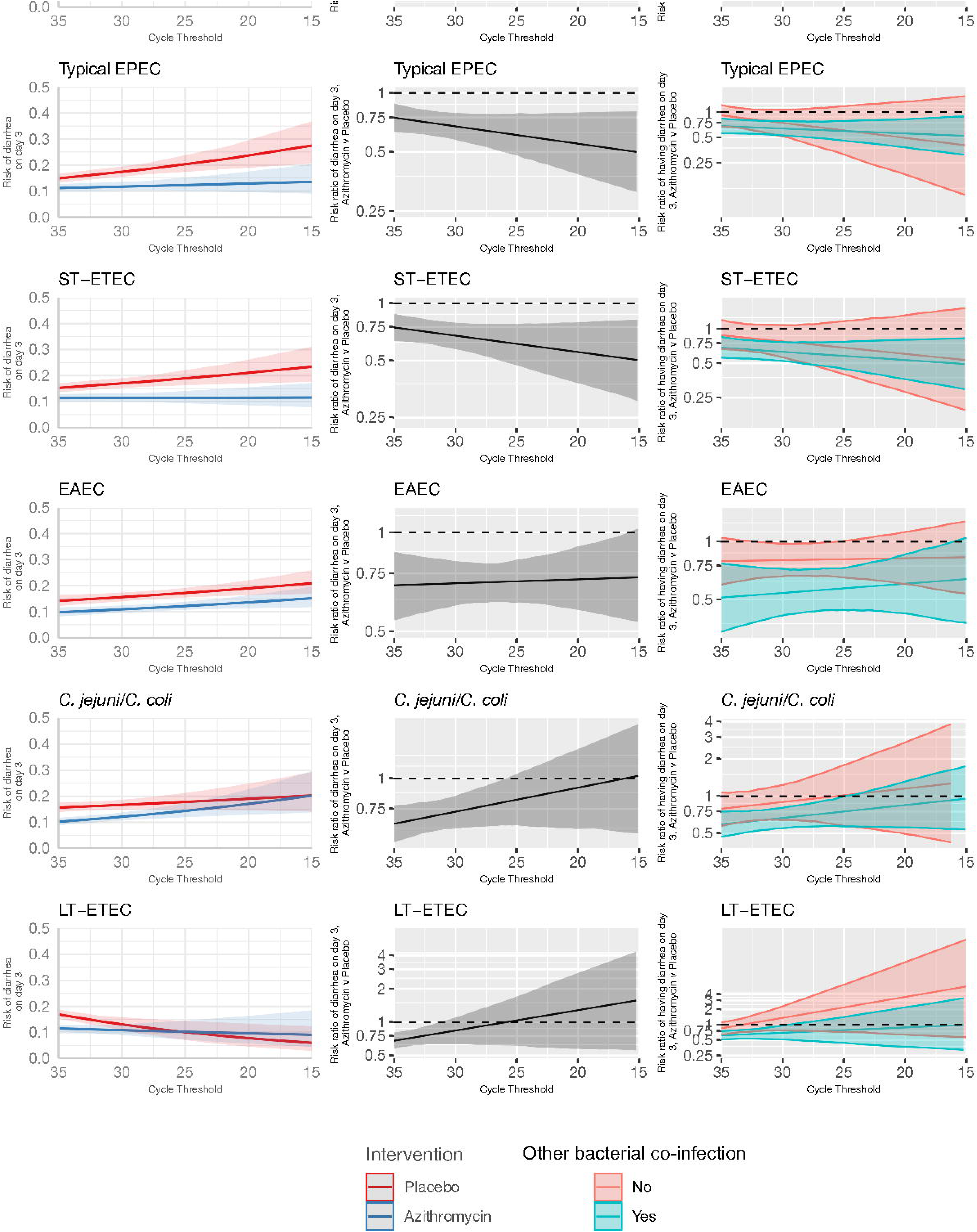
Modification of the effect of azithromycin on the risk of diarrhoea on day 3 by bacterial pathogen quantities detected. For each bacterial pathogen, **A:** the model-predicted risk of diarrhoea on day 3 by pathogen quantity and treatment arm (red = placebo; blue = azithromycin), conditional on sample collection at day 0 and no co- detections. **B:** the risk ratio for azithromycin compared to placebo for diarrhoea on day 3 by pathogen quantity adjusted for the day of diarrhoea on which the sample was collected and co-detection of other pathogens. **C:** the adjusted risk ratio for azithromycin compared to placebo for diarrhoea on day 3, stratified by the presence of a bacterial co- aetiology (green = yes; red = no). Pathogen quantity is specified by the cycle threshold value from qPCR (i.e., smaller cycle thresholds correspond to higher pathogen quantity detected). Bands on all plots indicate 95% confidence bands.

**Table 1.** Change in the effect of azithromycin on risk of diarrhoea on day 3 and risk rehospitalisation or death by day 90 for each log10 increase in pathogen quantity detected among children with watery diarrhoea in the AntiBiotics for Children with severe Diarrhoea (ABCD) Trial.

| Pathogen | Ratio of risk ratios (95% confidence interval) |  |
| --- | --- | --- |
|  | Diarrhoea on Day 3 | Rehospitalisation or death by day 90 |
| *Shigella*/EIEC | 0.87 (0.77, 0.97) | 0.93 (0.76, 1.13) |
| *V. cholerae* | 0.89 (0.68, 1.18) | 0.96 (0.45, 2.05) |
| tEPEC | 0.93 (0.85, 1.02) | 0.98 (0.84, 1.13) |
| *Cryptosporidium* | 0.94 (0.85, 1.03) | 0.87 (0.73, 1.04) |
| ST-ETEC | 0.94 (0.86, 1.02) | 1.00 (0.86, 1.15) |
| *Giardia* | 0.99 (0.87, 1.13) | 1.14 (0.95, 1.38) |
| *E. bienuesi* | 1.00 (0.74, 1.36) | 0.70 (0.43, 1.15) |
| Sapovirus | 1.00 (0.88, 1.12) | 0.93 (0.74, 1.16) |
| EAEC | 1.01 (0.93, 1.09) | 0.87 (0.76, 1.01) |
| Astrovirus | 1.06 (0.92, 1.21) | 0.99 (0.76, 1.29) |
| Norovirus GII | 1.06 (0.92, 1.21) | 1.04 (0.83, 1.30) |
| *Campylobacter jejuni/coli* | 1.08 (0.97, 1.19) | 1.01 (0.85, 1.21) |
| Adenovirus 40/41 | 1.09 (0.98, 1.23) | 1.13 (0.96, 1.33) |
| LT-ETEC | 1.10 (0.91, 1.35) | 1.17 (0.92, 1.50) |
| Rotavirus | 1.16 (1.06, 1.26) | 0.93 (0.78, 1.12) |

In contrast, there was no association between pathogen quantity detected and azithromycin treatment response for *Campylobacter jejuni/coli*, LT-ETEC, and EAEC. For *C. jejuni/coli*, the risk of diarrhoea on day 3 was higher with increasing pathogen quantities detected in both the azithromycin and placebo groups (Figure 1A). This resulted in risk ratios for diarrhoea on day 3 closer to the null at high *C. jejuni/coli* quantities. For LT-ETEC, the risk of diarrhoea on day 3 was very similar between azithromycin and placebo groups at high quantities detected, and for EAEC, the risk ratio for diarrhoea on day 3 was independent of pathogen quantity.

For viruses and parasites, the azithromycin treatment response was inversely associated or was not associated with pathogen quantity detected (Figure 2). For example, for rotavirus, norovirus GII, adenovirus 40/41, and astrovirus, the risk ratio of diarrhoea on day 3 was closer to the null with higher pathogen quantities detected.

**Figure 2.**
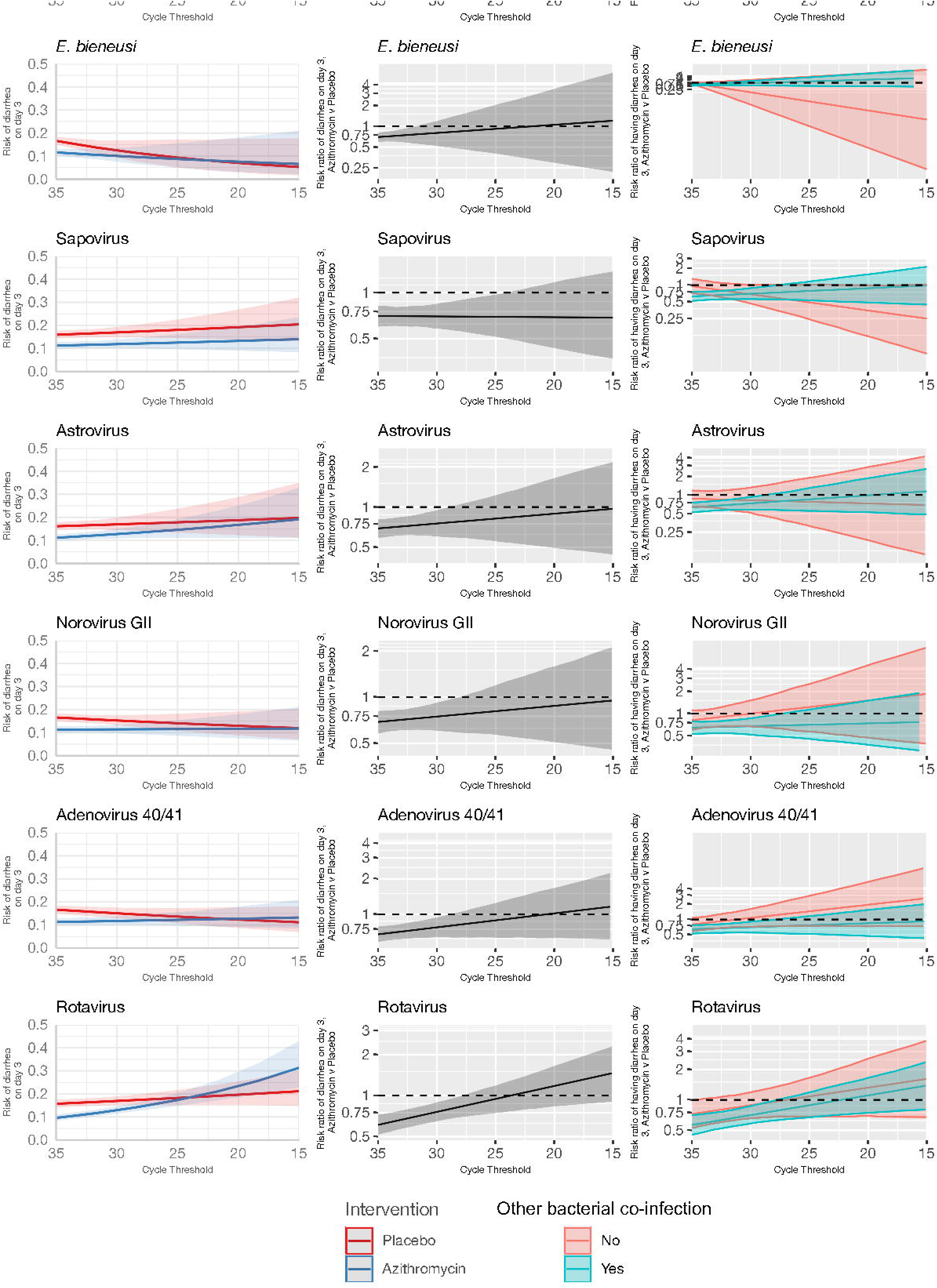
Modification of the effect of azithromycin on the risk of diarrhoea on day 3 by non-bacterial pathogen quantities detected. For each non-bacterial pathogen, **A:** the model-predicted risk of diarrhoea on day 3 by pathogen quantity and treatment arm (red = placebo; blue = azithromycin), conditional on sample collection at day 0 and no co-detections. **B:** the risk ratio for azithromycin compared to placebo for diarrhoea on day 3 by pathogen quantity adjusted for the day of diarrhoea on which the sample was collected and co-detection of other pathogens. **C:** the adjusted risk ratio for azithromycin compared to placebo for diarrhoea on day 3, stratified by the presence of a bacterial co- aetiology (green = yes; red = no). Pathogen quantity is specified by the cycle threshold value from qPCR (i.e., smaller cycle thresholds correspond to higher pathogen quantity detected). Bands on all plots indicate 95% confidence bands.

There was no consistent pattern when stratifying by presence of a bacterial co- aetiology. *Cryptosporidium* was an outlier; higher quantities detected were associated with a larger azithromycin treatment response. This was not explained by the presence of a bacterial co-aetiology. Rather, the azithromycin treatment response was more strongly associated with *Cryptosporidium* quantity for episodes without a bacterial co- aetiology.

These results were consistent for the combined outcome of rehospitalisation or death by day 90, but the associations were less precise and none of the effect heterogeneity was statistically significant (Figure 3 and Figure 4). The risk of death or hospitalisation was 4.5% overall, 3.6% in the azithromycin group and 5.3% in the placebo group. For episodes in which *Shigella* was detected at a higher quantity (Ct=25), azithromycin was associated with a 57% reduction (95% CI: 29%, 70%) in rehospitalisation or death, compared to a 44% reduction (95% CI: 37%, 54%) for episodes with *Shigella* detected at a lower Ct=30 quantity. The only discrepant result was for rotavirus, for which higher quantities were associated with slightly stronger azithromycin treatment response.

**Figure 3.**
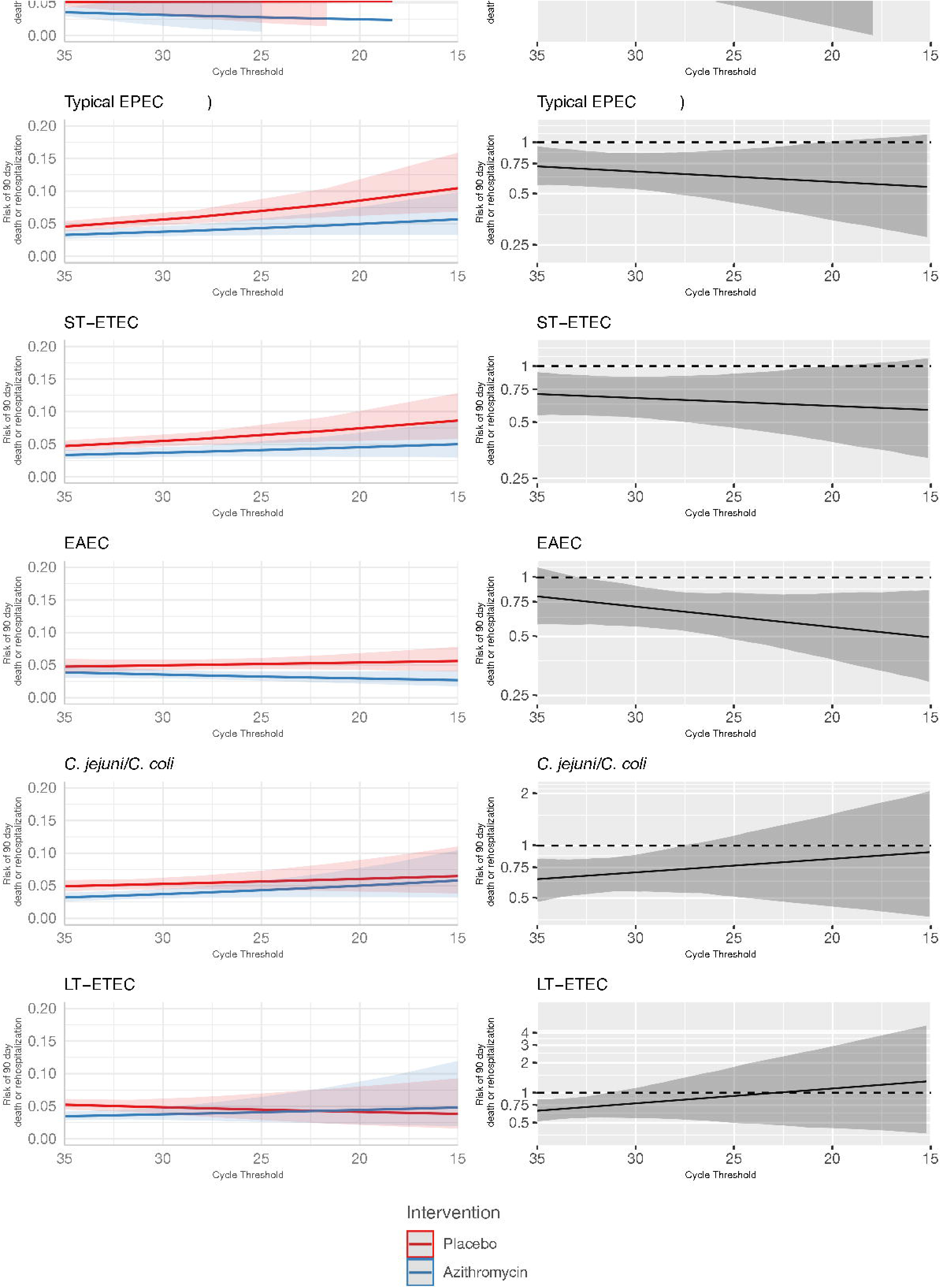
Modification of the effect of azithromycin on rehospitalisation or death by day 90 by bacterial pathogen quantities detected. For each bacterial pathogen, **A:** the model-predicted risk of rehospitalisation or death by pathogen quantity and treatment arm (red = placebo; blue = azithromycin), conditional on sample collection at day 0 and no co-detections. **B:** the risk ratio for azithromycin compared to placebo for rehospitalisation or death by pathogen quantity adjusted for the day of diarrhoea on which the sample was collected and co-detection of other pathogens. Pathogen quantity is specified by the cycle threshold value from qPCR (i.e., smaller cycle thresholds correspond to higher pathogen quantity detected). Bands on all plots indicate 95% confidence bands.

**Figure 4.**
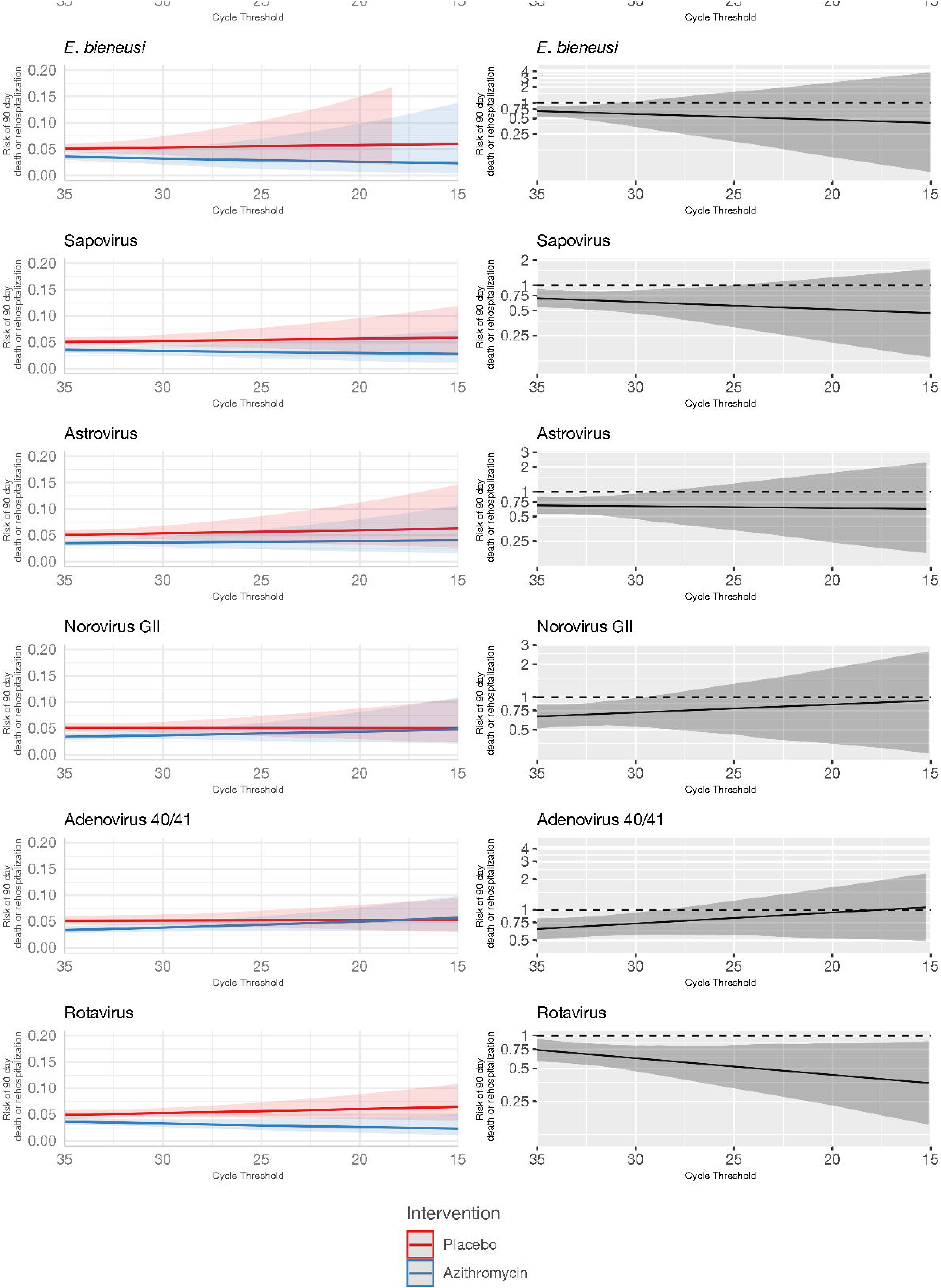
Modification of the effect of azithromycin on rehospitalisation or death by day 90 by non-bacterial pathogen quantities detected. For each non-bacterial pathogen, **A:** the model-predicted risk of rehospitalisation or death by pathogen quantity and treatment arm (red = placebo; blue = azithromycin), conditional on sample collection at day 0 and no co-detections. **B:** the risk ratio for azithromycin compared to placebo for rehospitalisation or death by pathogen quantity adjusted for the day of diarrhoea on which the sample was collected and co-detection of other pathogens. Pathogen quantity is specified by the cycle threshold value from qPCR (i.e., smaller cycle thresholds correspond to higher pathogen quantity detected). Bands on all plots indicate 95% confidence bands.

Using the azithromycin treatment response as a probe to attribute aetiology, we estimated that 4.14% of episodes could be attributed to *Shigella*, 2.95% could be attributed to ST-ETEC, 2.94% could be attributed to tEPEC, and 0.37% could be attributed to *Vibrio cholerae.* The population attributable fractions for *Campylobacter jejuni/coli*, EAEC, and LT-ETEC were zero.

The proportion of aetiology-specific and overall diarrhoea episodes that would be treated if treatment decisions were based on pathogen quantity cut-offs depended on the distribution of pathogen quantities among aetiology-specific episodes and the distribution of etiologies (Figure 5). For example, treatment of all episodes with tEPEC detected at a Ct of 25 or lower would result in 50% of episodes with tEPEC treated and 8% of episodes treated overall. In contrast, treatment of all episodes with *V. cholerae* detected at a Ct of 25 or less would result in only 16.6% of episodes with *V. cholerae* treated and 0.4% of episodes treated overall.

**Figure 5.**
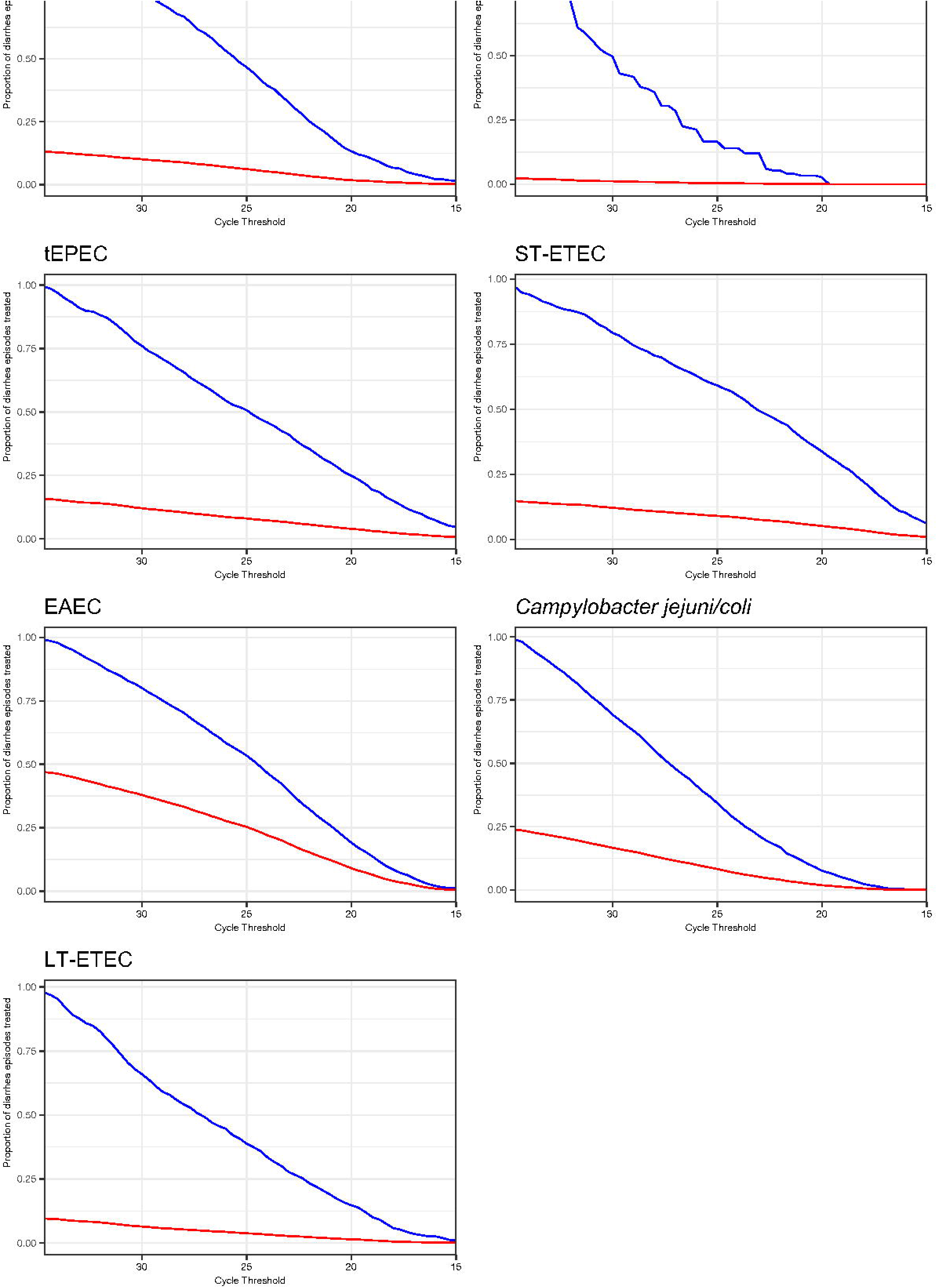
Prevalence of antibiotic treatment under decision rules that use cycle threshold cut-offs to define aetiology and assign treatment. The proportion of diarrhoea episodes overall (red line) and diarrhoea episodes with the pathogen detected at any quantity (blue line) that would be treated with antibiotics if a given cycle threshold were chosen as the cut-off to define aetiology and assign treatment.

## Discussion

In this reanalysis of the qPCR data from the ABCD study, the azithromycin treatment response at day 3 was stronger among episodes with higher detected pathogen quantities for some of the most common bacterial causes of diarrhoea: *Shigella*, *V. cholerae*, tEPEC, and ST-ETEC. This supports that these bacteria are likely the true aetiology of diarrhoea when detected at high pathogen quantities. While not statistically significant for all pathogens, the observed heterogeneity in effects by pathogen quantity was largely consistent with previous comparisons of qPCR data between diarrhoea cases and non-diarrhoeal controls from GEMS, which showed strong quantity dependent associations between all these bacterial pathogens and diarrhoea, with the exception of tEPEC; the highest quantities of tEPEC were only moderately associated with diarrhoea in GEMS (3). However, tEPEC was strongly associated with mortality in GEMS, providing additional support for tEPEC as an important cause of diarrhoea (15).

Our data suggest that the relationship between treatment response and bacterial pathogen quantity is approximately linear, making the assignment of a definitive cycle threshold cut-off for diarrhoeal aetiology difficult. Such a cut-off would be needed to define antibiotic treatment decision rules (i.e., treat if quantity < threshold and not if quantity > threshold) as well as to define case definitions for studies with pathogen- specific outcomes (e.g., *Shigella-*attributed diarrhea for a *Shigella* vaccine trial).

Pathogen quantity could however be used to guide treatment decisions, for example by choosing to treat at a quantity threshold corresponding to a clinically meaningful treatment benefit if a derivative of this assay becomes available as a point of care test. The choice of threshold should also consider the potential benefits of expanded use of azithromycin to treat episodes of bacterial diarrhoea in the context of the growing azithromycin resistance rates in key Gram-negative bacteria, including *Shigella* spp (16,17).

The lack of associations between pathogen quantity and azithromycin treatment response for *Campylobacter,* LT-ETEC, and EAEC were consistent with the weaker associations between pathogen quantities and diarrhoea observed for these pathogens in MAL-ED and GEMS (3,4). These prior studies of diarrhoea aetiology attributed relatively few diarrhoea episodes to these pathogens compared to their prevalence during diarrhoea, and our results corroborate that they may be relatively rare causes of diarrhoea in children in low-resource settings who have early and high levels of exposure. Sub-clinical carriage of these pathogens is frequent, complicating the assignment of a cut-off to attribute diarrhoea to this pathogen (18). Low qPCR cut-offs used to attribute *Campylobacter* diarrhoea based on MAL-ED and GEMS, for example used in the previous analysis of the ABCD study, have been criticized as potentially being poorly sensitive (7). However, the lack of treatment response even among episodes with high *Campylobacter* quantity support the use of a low Ct (i.e., high quantity) cut-off for attributing *Campylobacter a*etiology.

A key purpose of vaccine probe studies is to identify the proportion of disease attributable to the vaccine-targeted pathogen (19). In this “antibiotic probe” study analogue, we estimated population attributable fractions for the bacterial pathogens based on the azithromycin treatment response at observed pathogen quantities. The resulting relative ranking of pathogens matched that previously reported in GEMS and MAL-ED (20,21). However, the proportions attributable were smaller in magnitude, likely because azithromycin is not perfectly effective in preventing diarrhoea on day 3.

Importantly, no fraction of diarrhoea could be attributed to *Campylobacter*, EAEC, or LT-ETEC using the azithromycin treatment response as a probe given the inverse associations between quantities of these pathogens and the treatment response.

We also predominantly observed an inverse association between non-bacterial (virus and parasite) quantities and treatment response at day 3. High viral quantities were strongly associated with poor treatment outcomes, suggesting the true aetiology of these diarrhoeal cases was viral, as we would not expect an azithromycin treatment response in cases of viral diarrhoea. The observation of improved treatment outcomes at lower virus quantities, may be due to the increased likelihood of bacterial co-infection at lower virus quantities, such that the virus was carried at sub-clinical levels while the aetiology was truly bacterial.

Interestingly, higher quantities of rotavirus were associated with a small reduction in rates of death or hospitalisation at day 90. Dehydration and/or undernourishment were inclusion criteria for the ABCD study. Malnourishment can suppress the immune system; the WHO recommends routine treatment of children with severe acute malnutrition (SAM) with a broad-spectrum antibiotic and in some settings azithromycin administration for SAM has been reported to improve recovery and reduce mortality rates (18). Studies conducted in both Africa and Asia have reported that rotavirus infection is associated with under nutrition (22,23). It is possible that the improved azithromycin treatment outcome observed for higher quantities of rotavirus in this study, is due to the anti-inflammatory effects of azithromycin to treat malnutrition, which may have led to improved recovery from rotavirus infection (24). Further, improved outcomes could also be a result of azithromycin driven gut microbiota ablation, which has previously been shown to enhance humoral immunity and reduce severity of rotavirus diarrhoea (25) .

For parasites, high *Giardia* quantities were strongly associated with poor treatment outcomes. Interestingly, *Cryptosporidium* was unique amongst the non-bacterial pathogens such that high quantities were associated with improved treatment response at both day 3 and day 90. This improvement was not explained by presence of bacterial co-infections but has been observed previously in a case series of HIV-infected adults (26). The mechanism for improved treatment outcome is unclear, however it is possible that the immunomodulatory effects of azithromycin may again be contributing to improved recovery in *Cryptosporidium* infection.

This study has some limitations. We had no azithromycin susceptibility data from the bacterial pathogens tested for in this analysis, however phenotypic resistance in commensal *E.coli* from a subset of ABCD participants from all sites was 24% three months after enrolment. It is possible that azithromycin resistance amongst the Gram-negative bacterial pathogens reduced the effect of the azithromycin treatment.

Mechanistic studies of azithromycin have shown it exhibits immunomodulatory activity through the regulation of multiple inflammatory pathways (24). Our results may have therefore been confounded by children who did not have bacterial attributable diarrhoea but exhibited a treatment response due to the anti-inflammatory activity of azithromycin. In addition, this study of effect heterogeneity was underpowered since the primary ACBD trial was powered for population-level effects. While few estimates of heterogeneity were statistically significant, changes in the magnitude of the azithromycin effect remain informative. Finally, our dataset is derived from children enrolled in the ABCD study, which had restrictive enrolment criteria requiring all children to have dehydration or malnutrition. Therefore, the estimates of the proportions of children who would be treated under varying quantity cut-offs are not generalizable to all children with diarrhoea. Furthermore, the results may not be generalizable to high-income countries (HICs). *Campylobacter* spp are endemic and show high sub-clinical carriage in LMICs but not in HICs (27). It is feasible that there may be a clearer association between *Campylobacter* quantity and azithromycin treatment response in HIC settings where *Campylobacter* is not endemic.

In this reanalysis of qPCR diagnostic data from the ABCD trial, the relationship between bacterial quantity and treatment response for *Shigella*, V*. cholerae*, tEPEC, and ST- EAEC support that attribution of diarrhoea aetiology to these pathogens is more accurate as pathogen quantity increases and suggest that the use of a pathogen quantity cut-off to attribute aetiology is appropriate for observational and intervention studies of diarrhoea using molecular diagnostics. However, because the heterogeneity of treatment response was linear with pathogen quantity, the assignment of an aetiological qPCR cut-off remains context specific.

## Financial support

The ABCD trial and nested molecular diagnostics study was funded by the Bill & Melinda Gates Foundation (grant numbers OPP 1126331 and OPP 1179069). This study was funded by the Bill & Melinda Gates Foundation (INV-044317 to ETRM).

## Data Availability

All data produced in the present study are available upon reasonable request to the authors.

## Notes

### Competing Interest Statement

The authors have declared no competing interest.

### Clinical Trial

NCT03130114

### Clinical Protocols

https://cdn.clinicaltrials.gov/large-docs/14/NCT03130114/Prot_001.pdf

### Funding Statement

This study was funded by the Bill & Melinda Gates Foundation (grant numbers OPP 1126331 and OPP 1179069). This study was funded by the Bill & Melinda Gates Foundation (INV-044317 to ETRM).

### Author Declarations

Ethics approval was obtained from the World Health Organization Ethics Review Committee, Emory University, and from the participating countries.

## References

1. Diarrhoeal disease [Internet]. [cited 2024 Apr 24]. Available from: https://www.who.int/en/news-room/fact-sheets/detail/diarrhoeal-disease

2. WHO. World Health Organization Integrated Management of Childhood Illness ( IMCI ) Chart Booklet—Standard. Geneva (Switzerland): World Health Organization [Internet]. 2014 [cited 2024 Aug 26];(March):1–80. Available from: https://www.who.int/publications/m/item/integrated-management-of-childhood-illness---chart-booklet-(march-2014)

3. Liu J, Platts-Mills JA, Juma J, Kabir F, Nkeze J, Okoi C, et al. Use of quantitative molecular diagnostic methods to identify causes of diarrhoea in children: a reanalysis of the GEMS case-control study. The Lancet. 2016 Sep 24;388(10051):1291–301.

4. JA PM, J L, ET R. Use of quantitative molecular diagnostic methods to assess the aetiology, burden, and clinical characteristics of diarrhoea in children in low-resource settings: a reanalysis of the MAL-ED cohort study. Lancet Glob Health. 6:e1309–18.

5. Cohen AL, Platts-Mills JA, Nakamura T, Operario DJ, Antoni S, Mwenda JM, et al. Aetiology and incidence of diarrhoea requiring hospitalisation in children under 5 years of age in 28 low-income and middle-income countries: findings from the Global Pediatric Diarrhea Surveillance network. BMJ Glob Health. 2022 Sep 1;7(9):e009548.

6. Ahmed T, Chisti MJ, Rahman MW, Alam T, Ahmed D, Parvin I, et al. Effect of 3 Days of Oral Azithromycin on Young Children with Acute Diarrhea in Low-Resource Settings: A Randomized Clinical Trial. JAMA Netw Open. 2021 Dec 16;4(12).

7. Pavlinac PB, Platts-Mills JA, Liu J, Atlas HE, Gratz J, Operario D, et al. Azithromycin for Bacterial Watery Diarrhea: A Reanalysis of the AntiBiotics for Children With Severe Diarrhea (ABCD) Trial Incorporating Molecular Diagnostics. J Infect Dis. 2024 Apr 12;229(4).

8. JW S, RW F, SD P. Azithromycin and loperamide are comparable to levofloxacin and loperamide for the treatment of traveler’s diarrhea in United States military personnel in Turkey. Clin Infect Dis. 45:294–301.

9. D V, V T, M SP. Single oral dose of azithromycin versus 5 days of oral erythromycin or no antibiotic in treatment of Campylobacter enterocolitis in children: a prospective randomized assessor-blind study. J Pediatr Gastroenterol Nutr. 50:404–10.

10. WA K, C S, U D, MA S, ML B. Treatment of shigellosis: V. Comparison of azithromycin and ciprofloxacin. A double-blind, randomized, controlled trial. Ann Intern Med. 126:697– 703.

11. W B, A A. Randomized comparison of azithromycin versus cefixime for treatment of shigellosis in children. Pediatr Infect Dis J. 22:374–7.

12. Team AS. A double-blind placebo-controlled trial of azithromycin to reduce mortality and improve growth in high-risk young children with non-bloody diarrhoea in low resource settings: the Antibiotics for Children with Diarrhoea (ABCD) trial protocol. Trials. 21:71.

13. Pavlinac PB, Platts-Mills JA, Liu J, Atlas HE, Gratz J, Operario D, et al. Azithromycin for Bacterial Watery Diarrhea: A Reanalysis of the AntiBiotics for Children With Severe Diarrhea (ABCD) Trial Incorporating Molecular Diagnostics. J Infect Dis. 2024 Apr 12;229(4).

14. Platts-Mills JA, McQuade ETR. Assigning Pathogen Etiology for Childhood Diarrhea in High-Burden Settings: A Call for Innovative Approaches. J Infect Dis. 2023 Oct 1;228(7):814–7.

15. Levine MM, Nasrin D, Acácio S, Bassat Q, Powell H, Tennant SM, et al. Diarrhoeal disease and subsequent risk of death in infants and children residing in low-income and middle-income countries: analysis of the GEMS case-control study and 12-month GEMS- 1A follow-on study. Lancet Glob Health [Internet]. 2020 Feb 1 [cited 2024 Aug 26];8(2):e204. Available from: /pmc/articles/PMC7025325/

16. Nuzhat S, Das R, Das S, Islam S Bin, Palit P, Haque MA, et al. Antimicrobial resistance in shigellosis: A surveillance study among urban and rural children over 20 years in Bangladesh. PLoS One. 2022 Nov 1;17(11 November).

17. Baker KS, Dallman TJ, Ashton PM, Day M, Hughes G, Crook PD, et al. Intercontinental dissemination of azithromycin-resistant shigellosis through sexual transmission: A cross- sectional study. Lancet Infect Dis. 2015 Aug 1;15(8):913–21.

18. Houpt ER, Ferdous T, Ara R, Ibrahim M, Alam MM, Kabir M, et al. Clinical Outcomes of Drug-resistant Shigellosis Treated with Azithromycin in Bangladesh. Clinical Infectious Diseases. 2021 May 15;72(10):1793–8.

19. Feikin DR, Scott JAG, Gessner BD. Use of vaccines as probes to define disease burden. Lancet [Internet]. 2014 May 5 [cited 2024 Aug 26];383(9930):1762. Available from: /pmc/articles/PMC4682543/

20. Platts-Mills JA, Liu J, Rogawski ET, Kabir F, Lertsethtakarn P, Siguas M, et al. Use of quantitative molecular diagnostic methods to assess the aetiology, burden, and clinical characteristics of diarrhoea in children in low-resource settings: a reanalysis of the MAL- ED cohort study. Lancet Glob Health [Internet]. 2018 Dec 1 [cited 2024 Aug 26];6(12):e1309. Available from: /pmc/articles/PMC6227251/

21. Liu J, Platts-Mills JA, Juma J, Kabir F, Nkeze J, Okoi C, et al. Use of quantitative molecular diagnostic methods to identify causes of diarrhoea in children: a reanalysis of the GEMS case-control study. Lancet [Internet]. 2016 Sep 24 [cited 2024 Aug 26];388(10051):1291–301. Available from: https://pubmed.ncbi.nlm.nih.gov/27673470/

22. Das SK, Chisti MJ, Sarker MHR, Das J, Ahmed S, Shahunja KM, et al. Long-term impact of changing childhood malnutrition on rotavirus diarrhoea: Two decades of adjusted association with climate and socio-demographic factors from urban Bangladesh. PLoS One [Internet]. 2017 Sep 1 [cited 2024 Apr 24];12(9). Available from: https://pubmed.ncbi.nlm.nih.gov/28877163/

23. Gasparinho C, Piedade J, Mirante MC, Mendes C, Mayer C, Nery SV, et al. Characterization of rotavirus infection in children with acute gastroenteritis in Bengo province, Northwestern Angola, prior to vaccine introduction. PLoS One [Internet]. 2017 Apr 1 [cited 2024 Apr 24];12(4). Available from: https://pubmed.ncbi.nlm.nih.gov/28422995/

24. Zimmermann P, Ziesenitz VC, Curtis N, Ritz N. The immunomodulatory effects of macrolides-A systematic review of the underlying mechanisms. Front Immunol [Internet]. 2018 Mar 13 [cited 2024 Apr 24];9(MAR):326197. Available from: www.frontiersin.org

25. Uchiyama R, Chassaing B, Zhang B, Gewirtz AT. Antibiotic Treatment Suppresses Rotavirus Infection and Enhances Specific Humoral Immunity. J Infect Dis [Internet]. 2014 Jul 7 [cited 2024 Jun 18];210(2):171. Available from: /pmc/articles/PMC4399425/

26. Kadappu KK, Nagaraja M V., Rao P V., Shastry BA. Azithromycin as treatment for cryptosporidiosis in human immunodeficiency virus disease. J Postgrad Med [Internet]. 2002 Jul [cited 2024 Jun 18];48(3):179–81. Available from: https://pubmed.ncbi.nlm.nih.gov/12432190/

27. Epps SVR, Harvey RB, Hume ME, Phillips TD, Anderson RC, Nisbet DJ. Foodborne Campylobacter: Infections, Metabolism, Pathogenesis and Reservoirs. International Journal of Environmental Research and Public Health 2013, Vol 10, Pages 6292-6304 [Internet]. 2013 Nov 26 [cited 2024 Apr 24];10(12):6292–304. Available from: https://www.mdpi.com/1660-4601/10/12/6292/htm

