## Supplemental Materials for "Azithromycin Treatment Response as a Probe to Attribute Bacterial Aetiologies of Diarrhoea using Molecular Diagnostics: A Reanalysis of the AntiBiotics for Children with severe Diarrhoea (ABCD) Trial"

**Table S1**. P-values for the interaction term between treatment arm assignment and the quantity of pathogen detected when specified with a linear and quadratic term among children with watery diarrhea in the AntiBiotics for Children with severe Diarrhoea (ABCD) Trial.

| Pathogen | P-value of linear term from linear model | P-value of quadratic term from quadratic model |
|---|---|---|
| *Vibrio cholerae* | 0.4 | 0.9 |
| Rotavirus | 0.001 | 0.03 |
| *Shigella*/EIEC | 0.02 | 0.2 |
| ST-ETEC | 0.1 | 0.08 |
| Astrovirus | 0.4 | 0.7 |
| *Cryptosporidium* | 0.2 | 0.3 |
| Norovirus GII | 0.4 | 0.7 |
| Adenovirus 40/41 | 0.1 | 0.3 |
| *C. jejuni/C. coli* | 0.2 | 0.06 |
| tEPEC | 0.1 | 0.7 |
| Sapovirus | 0.9 | 0.02 |
| *E. bieneusi* | 0.5 | 0.6 |
| EAEC | 0.8 | 0.2 |
| *Giardia* | 0.8 | 0.2 |
| LT-ETEC | 0.1 | 0.3 |

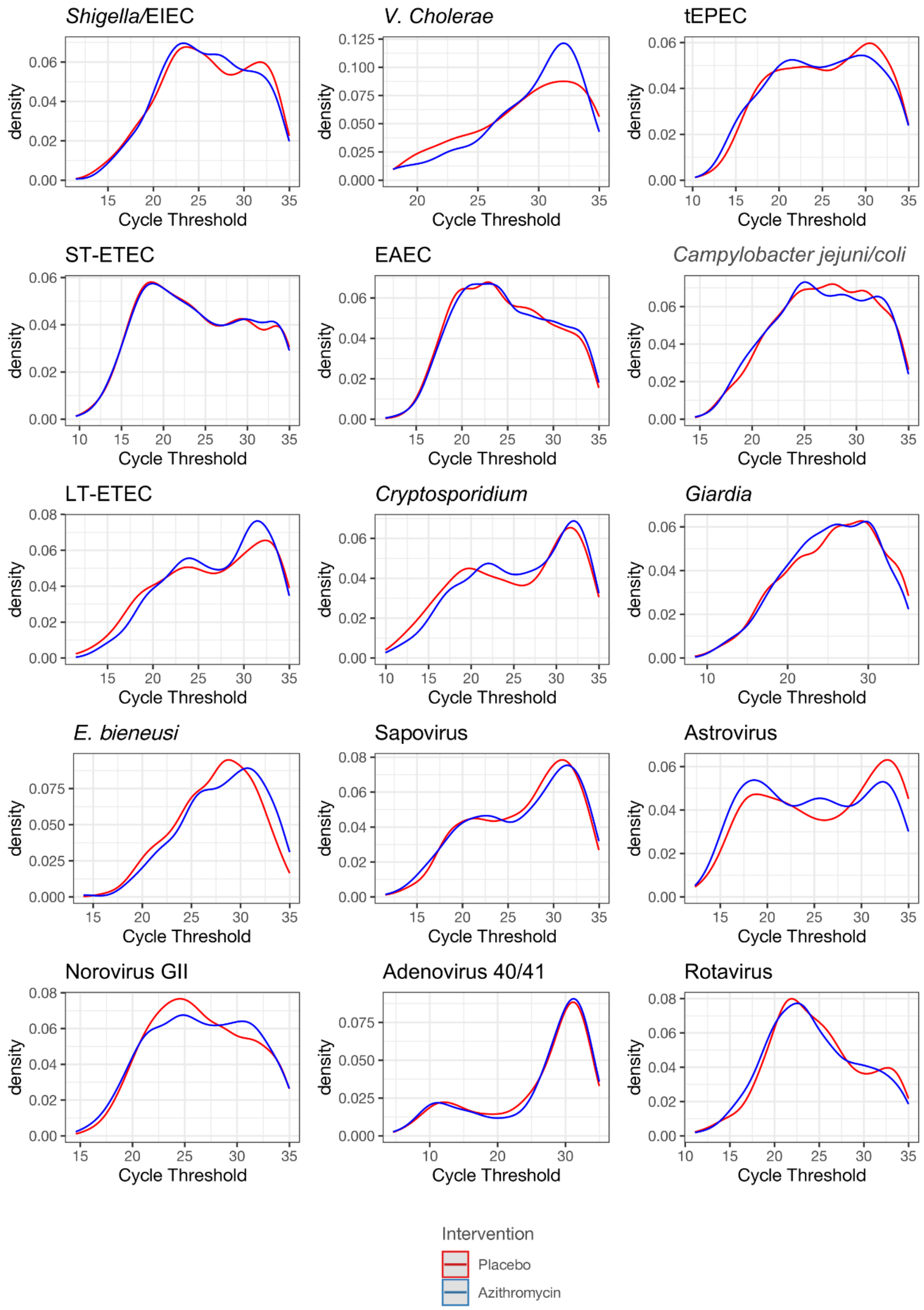


**Figure S1.** Density of pathogen quantities detected based on qPCR cycle threshold by intervention group (red = placebo; blue = azithromycin) in the ABCD trial.
